# A modelled analysis of the impact of COVID-19-related disruptions to HPV vaccination analysis

**DOI:** 10.1101/2023.03.07.23286911

**Authors:** Louiza S Velentzis, Megan A Smith, James Killen, Julia Brotherton, Rebecca Guy, Karen Canfell

**Author notes:** Corresponding author A/Prof Megan A Smith, PhD, The Daffodil Centre, The University of Sydney, a joint venture with Cancer Council NSW, Australia. Joint first authors (Louiza S Velentzis, Megan A Smith).

## Abstract

COVID-19 disrupted school attendance in many countries, delaying routine adolescent vaccination against human papillomavirus (HPV) in some settings. We used the ‘*Policy1-Cervix’* HPV model (natural history/vaccination/screening/HPV-related cancers), to estimate the impact on HPV-related cancers from disruptions to HPV vaccination in a high-income setting. Compared to no disruption (nonavalent vaccine uptake, age 12 [females:82.4%; males:75.5%] as in Australia), additional lifetime HPV-related cancer cases were calculated for three disruption scenarios affecting one birth cohort (2008): i) 1-year delay (no doses missed); ii) 1 to 7-year delay (slow catch-up); iii) no catch-up (herd effects only). A fourth scenario assumed no catch-up for two cohorts (2008,2009). We found a 1-year delay could result in ≤0.3% more HPV-related cancers (n=4) but the increase would be greater if catch-up was slower (5%; n=70), and especially if there was no catch-up (49%; n=750). Additional cancers for a single missed cohort were most commonly cervical (23%), oropharyngeal (males:20%) or anal (females:16%). In the worst-case scenario of two cohorts missing vaccination, ≤62% more HPV-related cancers would be diagnosed (n=1,892). In conclusion, providing catch-up of missed HPV vaccines is conducted, short-term delays in vaccinating adolescents are unlikely to have substantial long-term effects on cancer.

## Introduction

Australia was the first country to implement a fully-government funded National HPV Vaccination program. HPV vaccination has been routinely offered through schools, to girls aged 12–13 years since 2007 and to boys aged 12-13 years since 2013 (plus catch-up for females aged 14 to 26 years in 2007-2009 and for males age 14-15 in 2013-2014). In 2018, a three-dose schedule with the quadrivalent HPV vaccine which protects against four HPV types (6, 11, 16 and 18), was replaced by a two-dose schedule with the nonavalent HPV vaccine (HPV9), protecting against an additional five oncogenic HPV types (31, 33, 45, 52 and 58). Since 2017, free catch-up of vaccinations under the Australian National Immunization Program, including the HPV vaccine, is available to all people aged under 20 years and is administered in primary care.^1^ HPV course completion rates by age 15 were 79.8 % among girls and 77.0% among boys who reached the age of 15 years in 2019.^2^

Following the World Health Organization’s declaration of the COVID-19 pandemic in March 2020, strict national and state-level infection control measures were introduced in Australia. Social and physical restrictions affected essential health services including the delivery of school-based vaccination programs due to school closures from imposed lockdowns in 2020 and in some states in 2021.^3,4^ Globally, the World Health Organization reported the largest continued decline in overall vaccinations in the last three decades and a loss of over a quarter of the HPV vaccine coverage achieved in 2019.^5^

Infection with oncogenic HPV types is a major causal factor for the development of cervical cancer and for a fraction of cancers of the anus, oropharynx, penis, vagina, and vulva.^6^ Except for cervical screening, organized screening is not generally available for any of the other HPV-related cancers. A previous analysis of the burden of HPV-related disease in Australia estimated that 80% of the estimated 1,544 HPV-associated cancers in 2012 were attributable to types preventable by the quadrivalent vaccine (HPV16/18), with an additional 9% attributable to the five additional types prevented by the nonavalent vaccine.^7^

The aim of this study was to estimate the additional lifetime HPV-related cancer cases in men and women that could be caused by HPV vaccination delays or missed doses due to the pandemic by using a simulation model to project the long-term outcomes from different potential disruption scenarios, using Australia as an example. The study was designed prior to the emergence of actual 2020-21 vaccination disruption data, and hence, a number of potential disruption scenarios were considered to encompass a wide range of possibilities.

## Methods

### Model

We used Policy1-Cervix, an established modelling platform of dynamic HPV transmission, HPV natural history, cervical screening, treatment and cancer survival that has been validated across several settings. The dynamic HPV transmission component of Policy1-Cervix incorporates HPV vaccination and was used to estimate HPV incidence by age under scenarios of disruption to vaccination coverage compared to a no disruption scenario. The number of lifetime cervical cancer cases was subsequently estimated for the cohorts affected after explicitly modelling cervical screening (5-yearly HPV screening starting from age 25)^8^ to take into account that cervical screening may prevent some cancers that would otherwise have been prevented by vaccination. In addition, HPV incidence estimated by the dynamic HPV transmission model was used in a separate incidence-based model to project the lifetime number of non-cervical HPV-related cancers in both sexes (i.e., anal, oropharyngeal, penile, vaginal, vulvar), as previously described.^9^ Data included age- and sex-specific incidence of each cancer and proportion of cases attributable to vaccine-targeted HPV types (Supplementary Files 1A, 1B, 1C). In a sensitivity analysis, we also used an analogous incidence-based approach to also estimate the impact on cervical cancer, rather than explicitly allowing screening to prevent some of the cervical cancers which would arise due to missed vaccination. Further information on the Policy1-Cervix model can be found at https://www.policy1.org/models/cervix/documentation.

### Scenarios

A baseline scenario of uninterrupted HPV vaccination (status quo) assumed a 2-dose uptake of HPV9 at age 12 of 82.4% among females and 75.5% among males. Three disruption scenarios were modelled affecting the 2008 birth cohort who were aged 12 in 2020 (322,115 people). Scenario 1 assumed a fast catch-up where vaccination was delayed by one year (but no doses were entirely missed); scenario 2 assumed a slow catch-up where vaccination was delayed between 1 and 7 years with an equal proportion being caught up each year; and scenario 3 assumed there was no catch-up, and therefore any protection in the cohort would be due to herd effects only. A fourth scenario was also conducted which assumed disruption to HPV vaccination for two birth cohorts (cohort of 2008 and 2009) (644,230 people), with no catch-up.

For each scenario we estimated the number of HPV-associated cancer cases (from age 12 to 84 years) prevented by vaccination compared to an unvaccinated comparator cohort, and the additional number of cases under each disruption scenario compared to the no disruption scenario.

## Results

In the baseline scenario, we estimate 2,583 HPV-related cancer cases would be prevented in the 2008 cohort due to HPV vaccination, equating to 63% of all HPV-related cases (Table 1). The first disruption scenario (1-year delay) would result in 4 (0.3%) additional lifetime cancer cases and slightly fewer cases prevented overall (2,579). Under a slow vaccination catch-up strategy or in the worse cases - no vaccination catch-up for either one or two cohorts - an additional 70 (5%), 750 (49%) and 1892 (62%) cancer cases, respectively, were predicted to occur.

**Table 1:** Estimated number of HPV-related cancer cases and cases prevented for vaccinated and unvaccinated cohorts, and four vaccination disruption scenarios.

| Modelled scenarios | Affected Cohort (2008) |  |  |  |  | Affected Cohorts (2008 and 2009) |  |  |  |  |
| --- | --- | --- | --- | --- | --- | --- | --- | --- | --- | --- |
|  | Total Cases | Prevented Cases (Unvax Comparator) | Additional Cases (Compared to No disruption) | % Prevented (Unvax Comparator) | % Additional (Compared to No disruption) | Total Cases | Prevented Cases (Unvax Comparator) | Additional Cases (Compared to No disruption) | % Prevented (Unvax Comparator) | % Additional (Compared to No disruption) |
| Unvaxed | 3,923 |  |  |  |  | 7,847 |  |  |  |  |
| No disruption | 1,532 | 2,583 |  | 63% |  | 3,061 | 4,876 |  | 63% |  |
| S1: 1 y delay | 1,537 | 2,579 | 4 | 63% | 0.3% | 3,066 | 4,781 | 5 | 63% | 0.2% |
| S2: 1 to 7y delay | 1,603 | 2,513 | 70 | 61% | 5% | 3,133 | 4,714 | 72 | 62% | 2% |
| S3: no catch-up | 2,282 | 1,833 | 750 | 45% | 49% | 3,846 | 4,001 | 785 | 53% | 26% |
| S4: no catch-up (2 missed cohorts) | 2,503 | 1,613 | 970 | 39% | 63% | 4,954 | 2,893 | 1892 | 40% | 62% |

Abbreviations: vax: vaccine; unvaxed: unvaccinated (i.e. assuming no HPV vaccination in cohort(s)).
No disruption: status quo HPV vaccination in males and females at age 12; scenario 1: rapid catch-up, 1-year delay in HPV vaccine catch-up; scenario 2: slow catch-up, 1 to 7-year delay in
HPV vaccine catch-up; scenario 3: no HPV vaccine catch-up (herd effects only 2008 cohort); scenario 4: no HPV vaccine catch-up (herd effects only; 2008 and 2009 cohorts).

Table 2 shows a breakdown of HPV-related cancer cases developed over a lifetime by cancer type and sex for the disruption scenarios. In the worst-case scenarios of missed HPV vaccination, most of the additional cancers occurred in females, and the additional cancers were most commonly cervical and anal cancers (females) and oropharyngeal, followed by anal cancers (males). When combined across both sexes, the additional cancers were most commonly oropharyngeal (>80% of which were in males) and anal cancers (mostly in females). In the main analysis, we explicitly modelled primary HPV cervical screening as also occurring in these cohorts, which would allow the cervical screening program to compensate, to some extent, for cervical cancers not prevented via HPV vaccination. In the sensitivity analysis that used an incidence-based approach, the number of additional cervical cancer cases predicted was larger and cervical cancers would have comprised around 35% of the additional HPV-related cancer cases (rather than around 17-25%, when screening was modelled) (Supplementary File 1D). The percentage increase in cases was similar but somewhat higher (for example 54% increase in cases for one missed cohort, compared to 49% increase when screening was modelled).

**Table 2:** Estimated number of cancer cases in modelled scenarios according to sex and cancer type.

| Modelled scenarios | Total Cases (additional compared to no disruption) | Females, N (additional compared to no disruption) |  |  |  |  | Males, N (additional compared to no disruption) |  |  |
| --- | --- | --- | --- | --- | --- | --- | --- | --- | --- |
|  |  | Anal | Cervical | Oropharyngeal | Vaginal | Vulvar | Anal | Oropharyngeal | Penile |
| 2008 cohort |  |  |  |  |  |  |  |  |  |
| Unvaxed | 3,923 | 489 | 788 | 185 | 162 | 729 | 389 | 911 | 271 |
| No disruption | 1,532 | 100 | 62 | 86 | 61 | 580 | 74 | 423 | 146 |
| Scenario 1 | 1,537 (4) | 101 (1) | 63 (1) | 86 | 61 | 580 | 75 (1) | 424 (1) | 146 |
| Scenario 2 | 1,603 (70)* | 114 (14) | 74 (12) | 90 (4) | 64 (3) | 586 (6) | 85 (11) | 440 (17) | 150 (4) |
| Scenario 3 | 2,282 (750)* | 236 (136) | 250 (188) | 121 (35) | 96 (35) | 632 (52) | 175 (101) | 593 (170) | 180 (34) |
| 2008 and 2009 cohorts |  |  |  |  |  |  |  |  |  |
| Unvaxed | 7,847 | 978 | 1,576 | 370 | 324 | 1458 | 778 | 1822 | 542 |
| No disruption | 3,061 | 199 | 125 | 172 | 122 | 1160 | 148 | 845 | 292 |
| Scenario 4 |  |  |  |  |  |  |  |  |  |
| 2008 cohort | 2,503 (971)* | 276 (176) | 303 (241) | 131 (45) | 107 (46) | 647 (67) | 205 (131) | 644 (221) | 190 (44) |
| 2009 cohort | 2,451 (922) | 268 (169) | 285 (222) | 129 (43) | 104 (43) | 644 (64) | 199 (125) | 634 (212) | 188 (42) |
| Scenario 4 total | 4,954 (1,892)* | 544 (345) | 588 (463) | 260 (88) | 211 (89) | 1,291 (131) | 404 (256) | 1,278 (433) | 378 (86) |

\*The number of additional cases presented in the table do not always match the difference between case figures stated for individual scenarios in table 2, due to rounding. Unvaxed: assuming no HPV vaccination in cohort (s); No disruption: HPV vaccination in males and females at age 12 with coverage of 82.4% in females and 75.5% in males; scenario 1: rapid catch-up, 1-year delay; scenario 2: slow catch-up, 1 to 7-year delay; scenario 3: no catch-up (herd effects only 2008 cohort); scenario 4: no catch-up (herd effects only; 2008 and 2009 cohorts. Sum of cases may not add to 'total cases' due to rounding.

**Figure 1:**
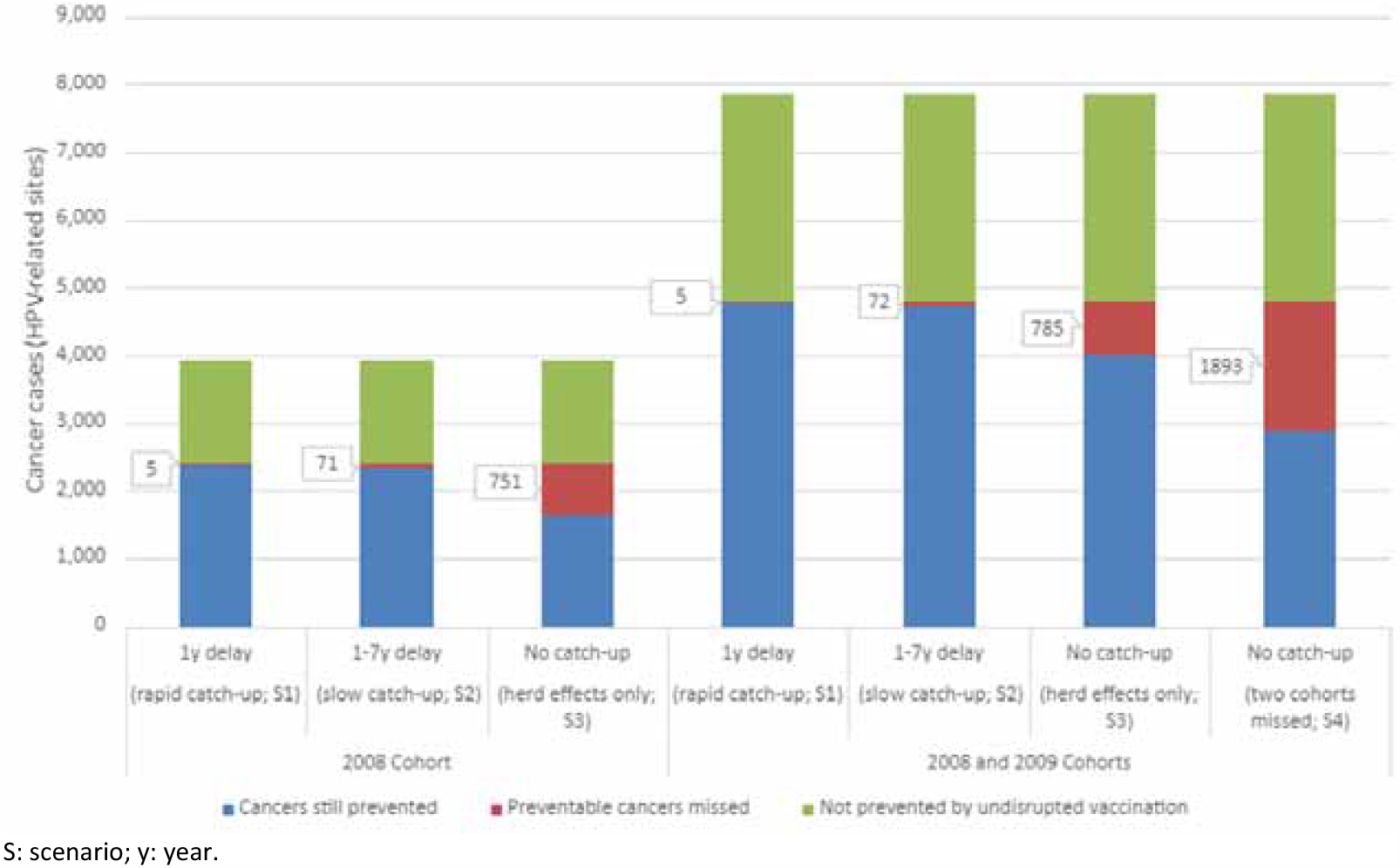
Estimated lifetime HPV-related cancer cases from two HPV vaccination catch-up scenarios [(1-year delay in vaccination (rapid); 1 to 7 year delay in vaccination (slow)] and two scenarios modelling the absence of vaccination catch-up, varying in the cohort affected (scenario 3 affecting the 2008 birth cohort; scenario 4 affectioning the 2008 and 2009 birth cohorts).

## Discussion

This is the first study to estimate the long-term potential impact of missed HPV vaccinations across HPV-induced cancers while accounting for natural history, sexual behaviour and herd immunity. We found that rapid catch-up of HPV vaccination in young adolescents within a year meant that even extreme disruptions where all vaccinations were delayed had a minimal long-term effect on cancer cases. A slower pace of recovery, however, or, in the worst case, missed vaccination in one or two cohorts was associated with a higher excess burden of cancers. While herd effects from vaccination still prevented close to half of HPV-related cancers, missing a single cohort entirely would result in an estimated 49% more HPV-related cancers in that cohort compared to no disruption.

In Australia, for adolescents aged 11 to <15 years who received their first dose in 2020, 74.7% of girls and 72.6% of boys, nationally, received their second dose in the same calendar year as their first dose.^2^ Corresponding figures for 2019, for the second dose, were 86.2% and 84.3%, for girls and boys respectively.^2^ The difference in numbers suggest vaccination delays, possibly due to COVID-19 effects including school closures during lockdowns, and decreased school attendance due to infections. Reports from other countries indicate that COVID-19 has delayed progress in achieving high coverage of HPV vaccination. For example, a US study found recovery after a marked decline in HPV dose administration in March to May 2020 (median 64%- 71%) was not fully achieved during June-September 2020.^10^

In November 2020 WHO released a global strategy calling for all countries to take action to achieve the elimination of cervical cancer as a public health problem within the next century.^11^ In the near future most lives will be saved globally through increased uptake of cervical screening and access to cancer treatment (Canfell 2020), but in the longer-term maintaining high coverage of HPV vaccination will be critical in reducing mortality and achieving global elimination.^12,13^ Globally, the timeframe to reach elimination is likely to be impacted by pandemic-induced delays in HPV vaccination delivery and also by global shortages of the HPV vaccine estimated to last until 2024 thus delaying the introduction of vaccination in low- and middle-income countries.^14^ In countries predicted to achieve cervical cancer elimination in the relatively short term, such as Australia, short delays of one year are not predicted to delay elimination.^8^

The strengths of this study include evaluating the impact of the COVID-19 pandemic across different HPV-related cancers, in both women and men and across a wide range of possibilities for vaccination disruption - two of which assumed a delay in HPV vaccination and two scenarios where the affected birth cohorts remained unvaccinated. Some limitations should also be noted. Even in the context of some cohorts missing vaccination entirely, herd effects still prevented a considerable proportion of HPV-related cancers overall; these effects would be smaller in settings with lower coverage.^15^ This is unlikely to affect our finding that catch-up within a year minimises the impact of disruptions, and so would likely strengthen the case for rapid catch-up in settings where herd effects are likely to be lower. The model also assumed that cervical screening would continue in birth cohorts offered HPV9, and, based on our sensitivity analysis that removed explicit modelling of screening, this prevented some cancers that would otherwise have occurred due to missed vaccination. In practice, screening may not continue in the same way in cohorts offered HPV9 as it would be much less cost-effective.^16^ As yet, no policy decision has been made for screening in these cohorts, however, and screening may in future be tailored based on individual vaccination status, rather than age-eligibility for vaccination. The model did not incorporate any potential changes in sexual behaviour which may affect estimates.

Our findings provide evidence that rapid catch-up of missed HPV vaccine doses would minimise the long-term impact of vaccination program disruptions, but delays would result in preventable HPV-related cancer cases in future.

## Data Availability

Information on data sources, microsimulation model parametrization, calibration to epidemiologic data, and calibration approach in line with good modeling practice can be found in the supplementary materials for this article and on-line (https://www.policy1.org/models/cervix/documentation). The current manuscript is a computational study involving modelling rather than direct analysis of primary datasets and no new datasets were created. The model codes for Policy1-cervix have been developed over decades, are proprietary property, and cannot be provided by the authors at this time. However, the Daffodil Centre is working to provide transparent and reproducible modelling code for forthcoming projects. Access to current code is possible only through appropriately resourced, supervised training at the Daffodil Centre.

## Data availability statement

Supporting information on data, data sources, model parametrization, calibration to epidemiologic data, and calibration approach in line with good modeling practice can be found partly in the supplementary materials for this article and partly on-line (https://www.policy1.org/models/cervix/documentation). The current manuscript is a computational study involving modelling rather than direct analysis of primary datasets and no new datasets have been created.

Code for the Policy1-cervix microsimulation model has been developed over decades, is proprietary property, and cannot be provided by the authors at this time. However, the Daffodil Centre is working to provide transparent and reproducible modelling code for forthcoming projects. Access to code is possible only through appropriately resourced supervision at the Daffodil Centre after submission and approval of a detailed proposal by the interested researcher. The corresponding author should be contacted in the first instance.

